# Strengthening implementation of integrated care for small and nutritionally at-risk infants under six months: pre-trial feasibility study

**DOI:** 10.1101/2024.03.27.24304963

**Authors:** Marie McGrath, Shimelis Girma, Melkamu Berhane, Mubarek Abera, Endashaw Hailu, Hatty Bathorp, Carlos Grijalva-Eternod, Mirkuzie Woldie, Alemseged Abdissa, Tsinuel Girma, Marko Kerac, Tracey Smythe

## Abstract

An integrated care pathway to manage small and nutritionally at-risk infants under 6 months (u6m) and their mothers (MAMI Care Pathway) is consistent with 2023 WHO malnutrition guidelines and is being tested in a randomised controlled trial (RCT) in Ethiopia. To optimize trial implementation, we investigated contextual fit with key local stakeholders. We used scenario-based interviews with 17 health workers and four district managers to explore perceived feasibility. Eighteen policymakers were also surveyed to explore policy coherence, demand, acceptability, evidence needs, opportunities and risks. The Bowen feasibility framework and an access to healthcare framework were adapted and applied. Health workers perceived the MAMI Care Pathway as feasible to implement with support to access services and provide care. The approach is acceptable, given consistency with national policies, local protocols and potential to improve routine care quality. Demand for more comprehensive, preventive and person-centred outpatient care was driven by concerns about unmet, hidden and costly care burden for health services and families. Inpatient care for all severe wasting treatment is inaccessible and unacceptable. Support for routine and expanded components, especially maternal mental health, is needed for successful implementation. Wider contextual factors may affect implementation fidelity and strength. Policymakers cautiously welcomed the approach, which resonates with national commitments, policies and plans but need evidence on how it can work within varied, complex contexts without further system overstretch. A responsive, pragmatic RCT will generate the most useful evidence for policymakers. Findings have informed trial preparation and implementation, including a realist evaluation to contextualise outcomes.

## 1.0 Introduction

In low- and middle-income countries (LMICs), an estimated 15.5% infants u6m are wasted, 17.4% are underweight, 19.9% are stunted (Kerac et al., 2021) and 14.7% are born low birth weight (LBW) (Okwaraji et al., 2024). Identified and described in many different but closely-related ways (McGrath et al., 2024), these infants are at increased risk of poor growth and development, sickness and death with long-term personal, societal, and inter-generational health consequences (Grey et al., 2021; Lawn et al., 2023; Victora et al., 2008; Cesar G. Victora et al., 2021). Babies may be born nutritionally vulnerable or become so in early life; being born too soon or too small is prevalent and problematic (Lawn et al., 2023) and incidence of stunting and wasting peaks in the first 6 months of life (C.G Victora et al., 2021). Wasting in early life increases risk of further wasting later (Mertens et al., 2020), contributing to the global burden of 47 million children who are wasted (Global Nutrition Report Stakeholder, 2020).

Ensuring children both survive and thrive is a global health priority reflected in the 2030 Sustainable Development Goals (SDGs) (United Nations, 2015). However, within life-course interventions to prevent and treat malnutrition (UNICEF, 2021, 2022; UNICEF et al., 2021), the care of at-risk infants u6m has long been overlooked. A 2010 report uncovered a high burden of wasting (Kerac et al., 2011), elevated mortality risk (Grijalva-Eternod et al., 2017), and widespread policy and service blind spots, particularly for outpatient care (ENN et al., 2010). The subsequent 2013 update of WHO malnutrition guidelines made transformational recommendations, most notable sanctioning the outpatient management for clinically stable malnourished infants u6m (WHO, 2013). To help put these guidelines into practice, an integrated care pathway approach, the MAMI Care Pathway, was collectively developed in a global expert peer collaboration by the MAMI Global Network (Grey et al., 2023; MAMI Global Network et al., 2021). Modelled on Integrated Management of Childhood Illness (IMCI) guidelines (World Health Organisation, 2014), it applies and expands on what exists to help embed continuity of quality care for small and nutritionally at-risk mother-infant pairs within health and nutrition services. Since then, the MAMI Global Network has facilitated implementation research and practical ‘learning by doing’ to help fill priority research gaps (Angood et al., 2015). MAMI Care Pathway principles and practices are reflected in the latest update of WHO recommendations on ‘management of infants u6m at risk of poor growth and development,’ a dedicated section of their new management of malnutrition guidelines (World Health Organisation, 2023).

Despite global policy progress, uptake of national uptake of WHO recommendations has been low and slow; a 2020 review found that only six out of 63 countries recommended outpatient care for infants u6m (Lelijveld et al., 2020). This includes Ethiopia, whose government pioneered and scaled outpatient care for severely malnourished children over 6 months of age (Ferew Lemma et al., 2012) but like other countries, has retained inpatient treatment for infants u6m due to lack of contextualised evidence on how to do outpatient care (Federal Ministry of Health Ethiopia, 2019). There are also system challenges; while integrated service delivery across nutrition and health is a strategic priority of the Ethiopian government, it has proved elusive partly due to inadequate mainstreaming of nutrition into relevant sectoral policies, strategies, programmes and operational plans (Federal Democratic Republic of Ethiopia, 2021).

To address these critical evidence gaps, the MAMI Care Pathway is undergoing testing in a four year (2020-2024) research partnership (MAMI RISE). This involves formative research, a two-arm, parallel group, randomised controlled trial (RCT) in outpatient health centres in Oromia Region, Ethiopia (https://www.isrctn.com/ISRCTN47300347), realist evaluation and economic evaluation. The RCT will embed a complex public health intervention within existing outpatient health services. Understanding how the MAMI Care Pathway may perform or not in outpatient health facilities is crucial to inform the trial implementation strategy (Pearson et al., 2020). Identifying factors that may impede or enable implementation can improve the conduct and quality of the RCT (Skivington et al., 2021) (Eldridge et al., 2016). Addressing realities in trial implementation also increases the potential to strengthen relevance and transferability to real world systems and services (O’Cathain et al., 2013).

The aim of this study was to improve future MAMI Care Pathway implementation. Objectives towards this were to: explore feasibility including acceptability as perceived by health workers and national policymakers in Ethiopia, and to contribute evidence on pre-trial feasibility studies as a means of strengthening contextual fit and improving usability of generated evidence.

**Key messages**

- Evidence gaps prevail in how to manage small and nutritionally at-risk infants under 6 months in outpatient care
- The MAMI Care Pathway integrated approach is likely feasible to implement during a randomised control trial in outpatient clinics in Ethiopia if contextualised support is provided to enable accessible, quality care.
- Training, staffing, space and supplies are needed to address existing constraints and for new components.
- Maternal services, especially maternal mental health, are lacking.
- Research co-creation and partnership strengthens feasibility potential.
- Pre-trial feasibility studies strengthen trial implementation readiness.
- A context responsive RCT will generate the most useful evidence for national policymakers.

## 2.0 Methods

We report according to the Standards for Reporting Qualitative Research (SRQR) (O’Brien et al., 2014) and consolidated criteria for reporting qualitative research (COREQ) (Tong et al., 2007). Reflexivity is integrated through methods, results and discussion (Olaghere, 2022).

### 2.1 Setting

This study took place in Jimma Zone and in Meta Woreda (district), East Hararghe Zone, Oromia region, Ethiopia. Oromia region has a high burden of child malnutrition; stunting, wasting and underweight prevalence (0-59 months), 35.3%, 4.3% and 16.3% respectively (Ethiopian Public Health Institute (EPHI) [Ethiopia] & ICF, 2021). The RCT is located in Jimma Zone and Deder Woreda. In Jimma Zone, health workers for this study were selected from centres that would not be included in the RCT to avoid potential bias. Since all health facilities in Deder Woreda are included in the trial, the neighbouring Meta Woreda was selected (similar population and service profile to Deder). Policymakers and key stakeholders were nationally selected from across Ethiopia.

### 2.2 Participants

Primary health care units (health centres staffed by clinic nurses and health posts operated through health extension workers (HEWs)) provide primary care services in Ethiopia (Ministry of Health Ethiopia, 2021). Purposive sampling identified health workers for interview with experience of delivering care to at-risk infants u6m and mothers in inpatient, outpatient and community health services. We prioritised staff cadre who would deliver care during the RCT (outpatient clinic nurses). Mothers and families were not interviewed; health workers were asked how feasible the approach would be for mothers. Purposive and referral sampling identified policymakers involved in health and nutrition policy development and service planning for survey and/or interview.

### 2.3 Data collection

Health worker interviews took place September to October 2021. Policymaker survey/interviews took place September 2021 to March 2022. The RCT commenced 22^nd^ August 2022. A UK-based researcher (BSc, part-time PhD candidate, Charity Technical Director, female) led the study in collaboration with the MAMI RISE Team. An experienced Jimma University field researcher (BSc, clinical nurse, lecturer Jimma University, male) conducted the health worker interviews. Both researchers completed research ethics and good clinical practice training. The lead researcher administered the online survey and conducted remote interviews with policymakers. In-country researchers provided administrative and logistical support. Both the lead researcher and field researcher are also members of the MAMI RISE Research Team who are conducting the RCT.

We sampled 42 individuals in total (Figure 1). We conducted in-depth interviews with 21 health workers (clinic nurses, HEWs, district (woreda) managers), 11 from urban clinics/hospital in Jimma town and 10 in Meta Woreda from rural and urban settings. One clinic nurse initially consented to participate but could not (due to sickness). Eighteen senior stakeholders completed the online survey: five Ministry of Health (MOH) representatives, one United Nations (UN) agency, three donors, eight international non-governmental organisations (NGOs) and one consultancy organisation. Nine interviews were conducted (six had completed the survey and three had not), comprising five MOH personnel, one UN agency, one INGO, one donor and one consultancy organisation. Two UN agency representatives invited for interview were not available (overall 81% response rate).

**Figure 1:**
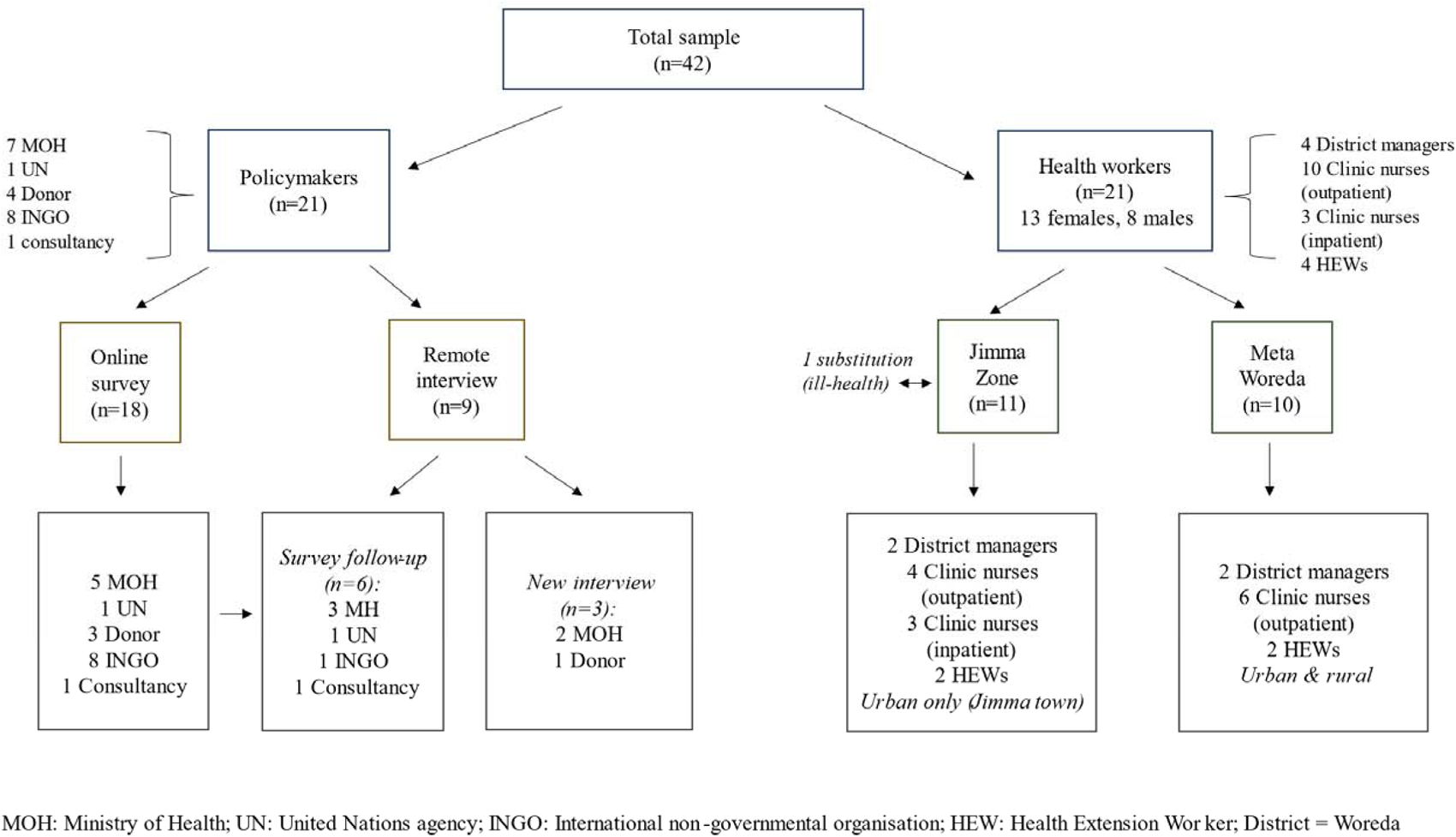
Sampling frame and size

**Figure 2:**
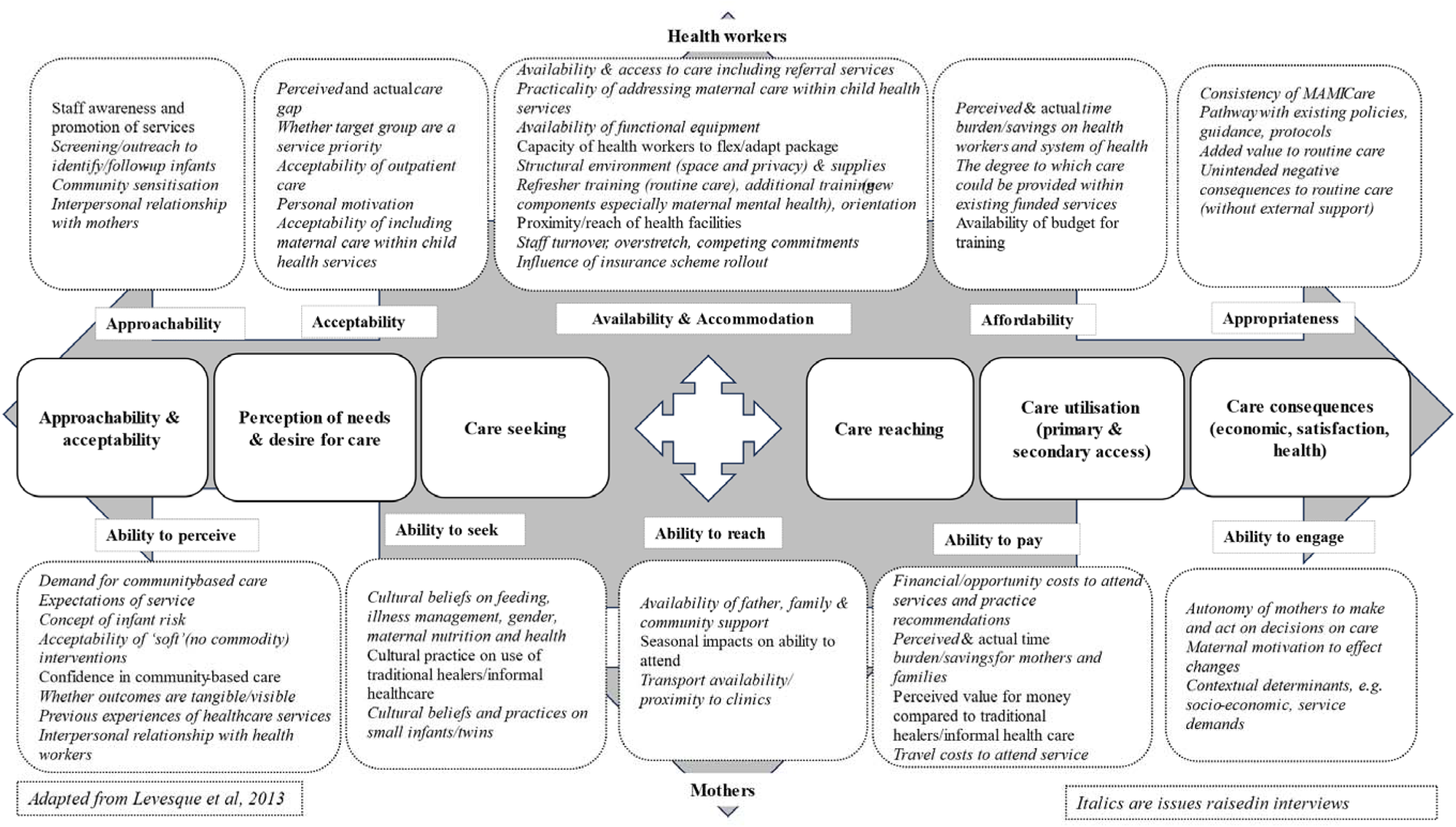
Abilily lo Access MAM] Care Framework

#### Health worker interviews

All health workers participated in a two day in-person orientation on the MAMI Care Pathway and implementation materials within two weeks of their interview. The field researcher helped deliver the workshop in both settings. Stakeholder-specific interview guides were developed in English by the lead researcher in consultation with the field researcher and MAMI RISE Team. Guides were translated into Afan Oromo and Amharic, and locally cross-checked and piloted with the research team and a clinic nurse (Jimma).

Health worker interviews were conducted by the field researcher in September (Meta Woreda) and October (Jimma Zone) 2021. Having already met interviewees during the workshop, the field researcher re-introduced himself, his clinical background and his role in this study. Study information was presented verbally and on paper. The field researcher provided information, consented and conducted interviews in Afan Oromo or Amharic as preferred. Interviews were scheduled at the participant’s convenience and conducted privately at their place of work. Locally appropriate compensation for 1.5 hours interview time was provided (10 USD mobile data credit). Interviews were recorded and transcribed directly from Amharic or Afan Oromo into English by the field researcher. Transcripts were not returned for interviewee review.

We used a scenario-based approach in one-to-one interviews to explore perceived feasibility as follows:

- Health workers were introduced to two practical mother-infant case scenarios and invited to talk through how they would apply the *MAMI Care Pathway* materials to each. They were encouraged to share their logic and process as they worked through the scenarios. We explored how useful they thought the questions would be in helping understand the situation of the mother and her baby, how comfortable they would feel in asking them, and how this compared to their usual practice. All were then asked to score the materials on a scale of 1-5 regarding usefulness, acceptability, practicality and improved care.
- For district managers, we used the MAMI Care Pathway Framework ‘who, what, where’ schematic to identify where MAMI Care Pathway activities already happen (or not) in the health facilities they manage (Supplementary Appendix 1).
- For both health workers and district managers, we then asked a series of open questions on the appropriateness of applying the MAMI Care Pathway approach in Ethiopia including ‘fit’ with staff roles and responsibilities, perceived acceptability to mothers, what training or support health workers would need for implementation, and what they see as the main implementation challenges in outpatient health facilities. We explored maternal mental health specifically, anticipating challenges informed by earlier formative work and team consultation.
- Finally, for all we asked who we should talk to about the research to make it work better and invited further ideas and questions.

#### Senior stakeholder survey and interviews

An online survey (Online Surveys v2, Joint Information Systems Committee) was distributed in English by the lead researcher through email to key contacts and networks across specialties and institutions in Ethiopia (survey live 29^th^ September 2021 to 30^th^ March 2022). The survey asked the extent to which the MAMI Care Pathway approach was needed (demand), its consistency with national health and nutrition policy guidance and plans (consistency), and how possible and how appropriate it was to implement in outpatient settings in Ethiopia including gaps or inconsistencies in the materials and potential opportunities, barriers and harms (acceptability). We also asked what supporting evidence or proof of effectiveness was needed before considering its use in Ethiopia (evidencing policy) and any opportunities in that regard (opportunities).

A sample of survey respondents was purposively selected for follow-up interview. A conversational style was used to elaborate on survey responses (n=6) or as an alternative to the online survey (n=3), guided by the survey structure. Interviewers were encouraged to ask questions and make suggestions regarding the RCT.

Private online interviews were conducted by the lead researcher in English between 25^th^ October 2021 and 22^nd^ February 2022. The lead researcher introduced herself, her background (25 years in international nutrition), role in the MAMI RISE Team and lead role in the feasibility study. Interviews were transcribed by the lead researcher aided by automated transcription (otter.ai).

### 2.4 Data analysis

All coding and data analysis was conducted by the lead researcher in the UK. Emerging findings were reviewed with a second researcher (TS) and discussed with the MAMI RISE Team, to inform conclusions and identify implications for the trial.

Health worker interview data were managed and coded using NVivo, v10 software, QSR International, 2012 (K Jackson & Bazeley, 2019). Free coding was first conducted to inductively explore the data, guided by and mapped to an adapted ‘access to MAMI care’ framework (Levesque et al., 2013). Deductive thematic analysis was then performed using an adapted version of the Bowen framework (Bowen et al., 2009) to investigate perceived feasibility and relevant phrases were coded (see Table 1). We applied four of the eight themes of the Bowen framework (acceptability, demand, implementation and practicality). We did not categorise ‘adaptation’ (modifications), ‘integration’ (system changes) or ‘expansion’ (to different populations/settings) as distinct Bowen themes, since these will be examined in the RCT realist evaluation. However, these dimensions are inherent to the MAMI Care Pathway approach (an integrated care model) that is being trialled within existing services in Ethiopia that involves ‘adaptations’ in an ‘expanded’ setting (inpatient to outpatient) and to an ‘expanded’ population (from severely wasted to a mix of vulnerable mother-infant pairs).

**Table 1:** Adapted Bowen Framework to investigate perceived feasibility of MAMI Care Pathway approach.

| Area of focus | This study explored | Sample outcomes of interest |
| --- | --- | --- |
| Acceptability | The extent to which the intervention is judged to be attractive, appropriate, suitable, satisfying to health workers and to mothers | Consistency with existing policies and services<br>Consistency with planned developments/other initiatives<br>Novelty value<br>Staff workload at health facility<br>Job satisfaction<br>Cultural/social acceptability of the MAMI approach<br>Key decision makers/influencers<br>Possible reasons of how and why it would be acceptable or could be made so<br>*MAMI Care Pathway materials:* format, content, images. nature of questions (content, how they are asked, by whom, cultural acceptability)<br>Rating score |
| Demand | How much demand is likely to exist for MAMI outpatient care from health workers and mothers | Perceived burden of at-risk infants u6m<br>Gaps in availability, accessibility in current care<br>Staff desire, (dis)satisfaction<br>Mother/family desire, (dis)satisfaction<br>Consequences of shortfalls in current care<br>Added value to existing services<br>Possible factors affecting demand and differences in demand between different actors in the system and consequences of current shortfalls in care provision<br>*MAMI Care Pathway materials:* Complement/address gaps in existing job aids/guides/resource materials/missing materials |
| Implementation<sup>†</sup> | The extent to which MAMI Care **could be** successfully delivered by existing staff and services within outpatient health facilities | Stated as “possible” to imple,ment<br>Consistent with current or planned policies and protocols<br>Consistent with staff roles & responsibilities<br>Existence of necessary health and nutrition services |
|  |  | Existence of referral pathways to and from specialist services/community-based providers<br>Staff/mother's time<br>Implementation strength of routine care<br>Staff competencies and capacity<br>*MAMI Care Pathway materials:*<br>Consistency/added value compared to existing materials |
| Practicality<sup>†</sup> | The extent to which MAMI Care **would be** successfully delivered using existing means, resources and circumstances and without outside intervention within outpatient health facilities<br>*Identify what may be needed to help accommodate the intervention during RCT* | Functionality of referral pathways<br>Implementation strength of routine care<br>Gaps in service provision<br>External inputs needed for service, e.g. training, equipment, staff<br>External inputs needed for mothers/families, e.g. expenses, time<br>Day-to-day operational factors<br>Wider contextual issues<br>*MAMI Care Pathway materials:*<br>Corrections/adaptations identified |

<sup>†</sup>*Expansion and integration are inherent in the MAMI Care Pathway RCT that is testing an approach being expanded to a new setting (outpatient care) and to new population (beyond severely wasted infants) integrated within outpatient health facilities.*

Therefore, considerations around these aspects that emerged in interviews are embedded within acceptability, demand, implementation and practicality themes according to how they were expressed. ‘Limited-efficacy testing’ was not applicable to this type of feasibility study. Summary statistics (percentage) using MS Excel were calculated to compare the ratings score given by health workers to the MAMI Care Pathway materials.

Senior stakeholder survey and interview data were inductively and deductively analysed and coded to demand, consistency, acceptability, evidencing policy and opportunities, using MS Excel. A narrative synthesis of findings was undertaken and illustrated by anonymised quotes.

### 2.5 Ethical clearance

The study secured ethical approval of Jimma University Institutional Review Board (IRB) and LSHTM Research Ethics Committee. Permissions from local authorities to conduct the study were secured by the local team. For health workers, informed written consent was secured by the field researcher immediately before each interview. For the survey, online informed consent was mandatory to proceed. Consent to approach for follow-up interview was sought as part of the online survey and confirmation sought by email in advance of the interview or confirmed verbally at the beginning of the interview, along with consent to record.

## 3.0 Results

### 3.1 Characteristics of population

Amongst health workers, professional experience and training varied. Eight reported BSc qualification and one MSc (manager). Over half (n=12) had minimum 6 years’ practical experience, of whom 6 had more than 10 years. Ten clinic nurses/HEWs had received training on Integrated Management of Newborn and Childhood Illness (IMNCI), breastfeeding counselling, acute malnutrition management and infant and young child feeding (IYCF) to varying degrees; seven reported no further training since primary qualification. Most had not received recent training. All health workers performed well with the scenarios, correctly classifying and accessing support materials during the interview. Professional qualification and experience were not assessed for policymakers.

### 3.2 Perceived feasibility

Perceptions amongst health workers and district managers are described by feasibility theme below. We found considerable interaction and fluidity across and within Bowen themes, which limited strict categorisation by theme or person profile. Summary findings by theme and implications for RCT planning and implementation are provided in Table 2. Findings were also mapped against the adapted access MAMI care framework (see Figure 3).

**Table 2:**
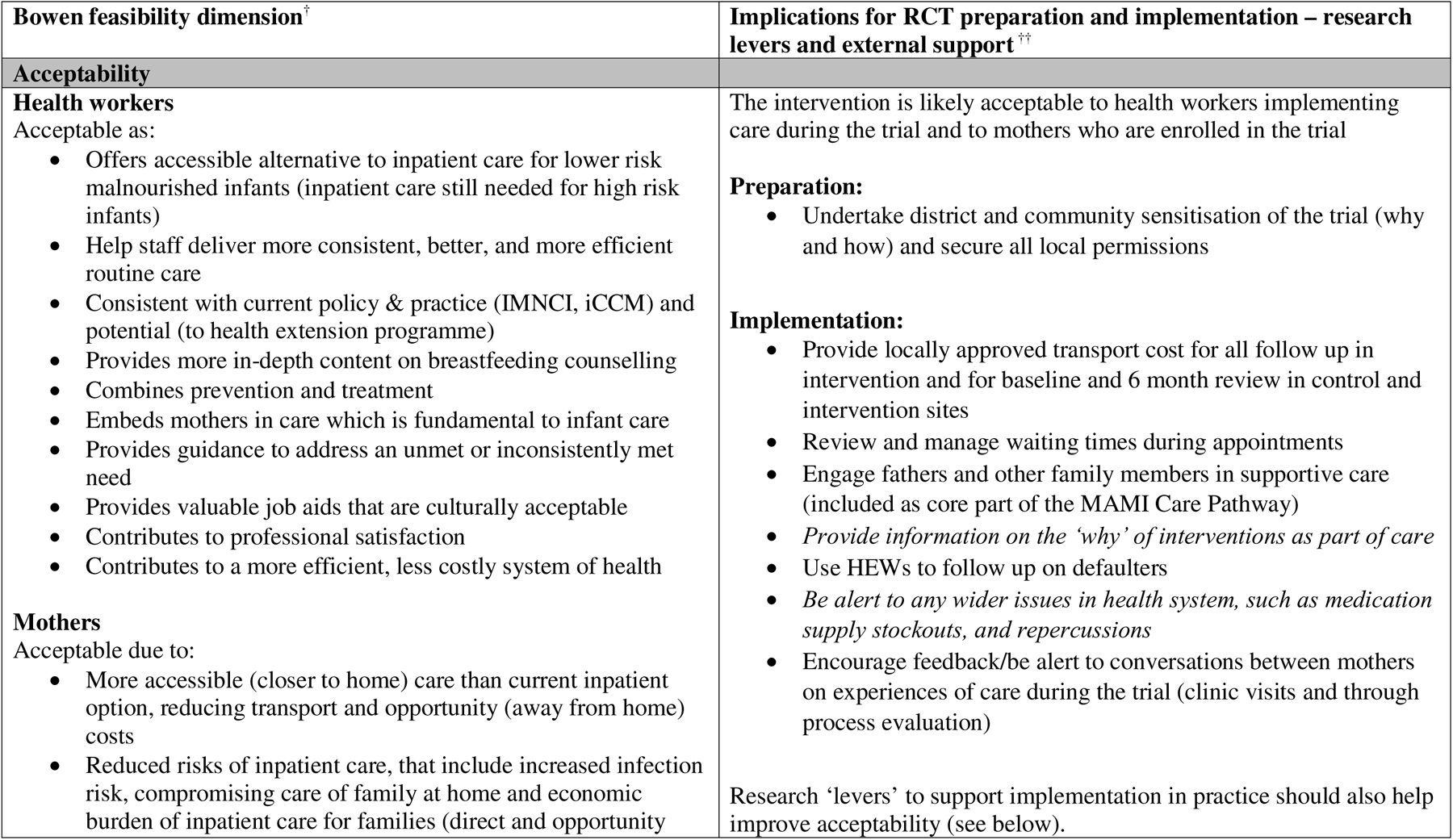
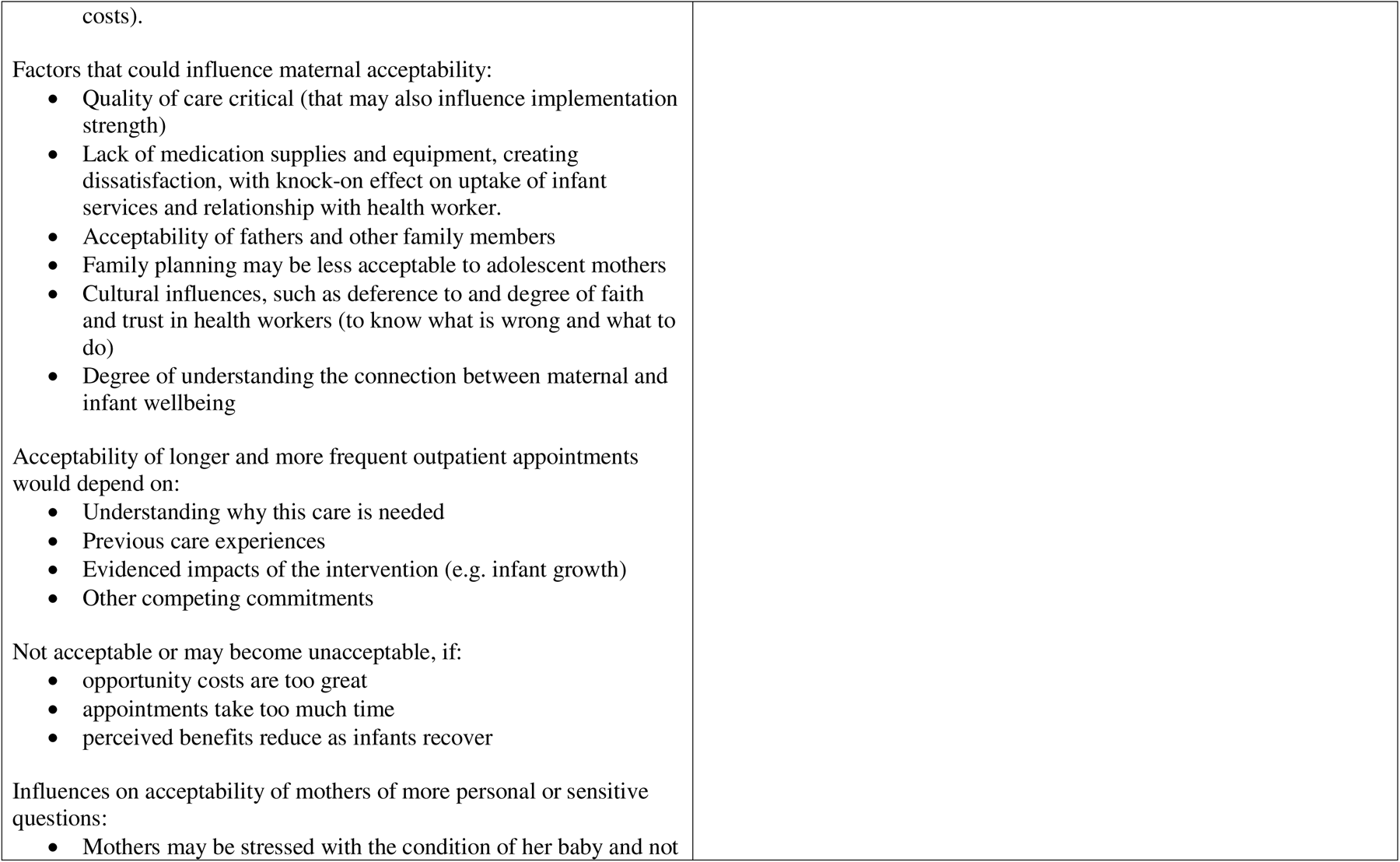
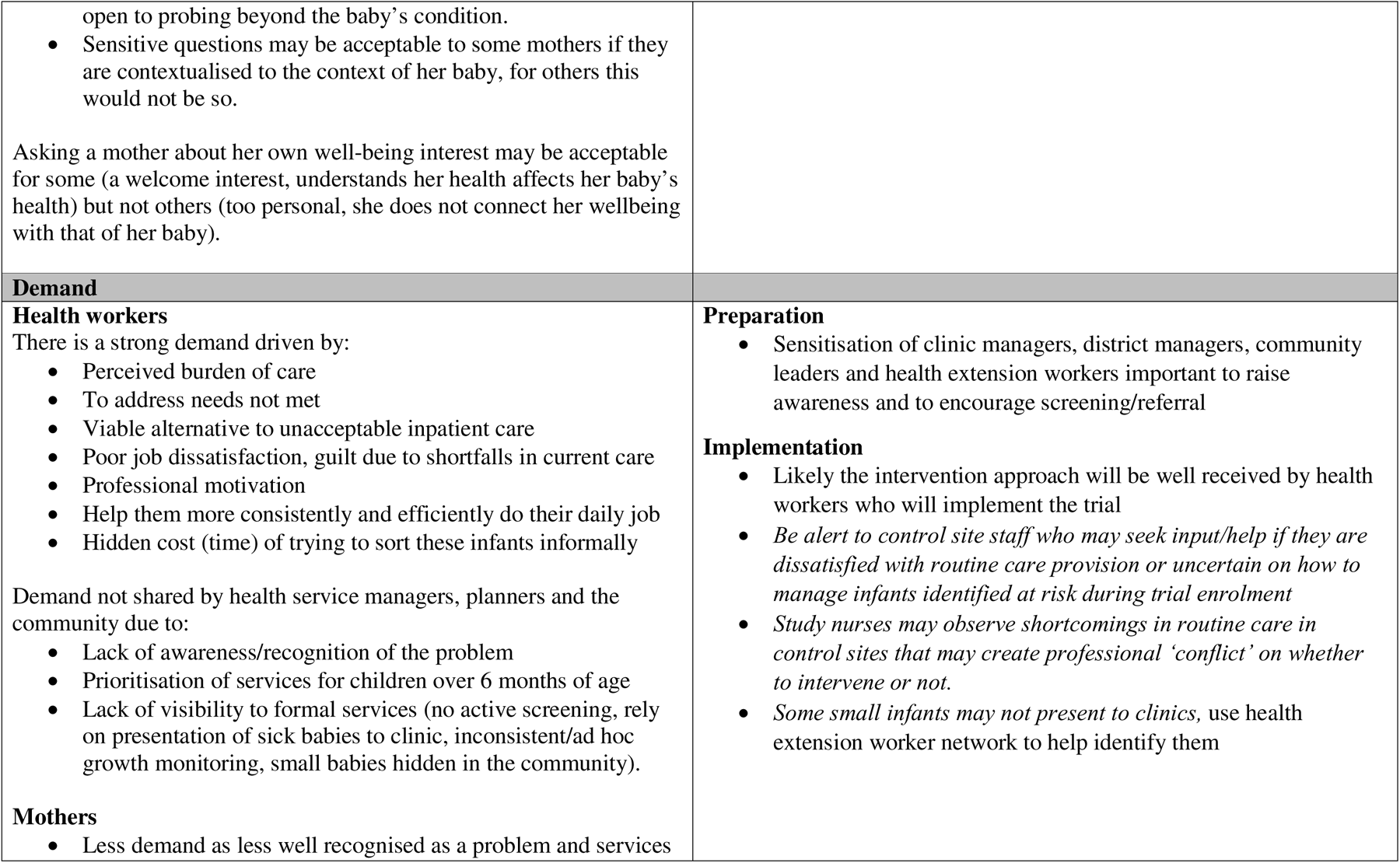
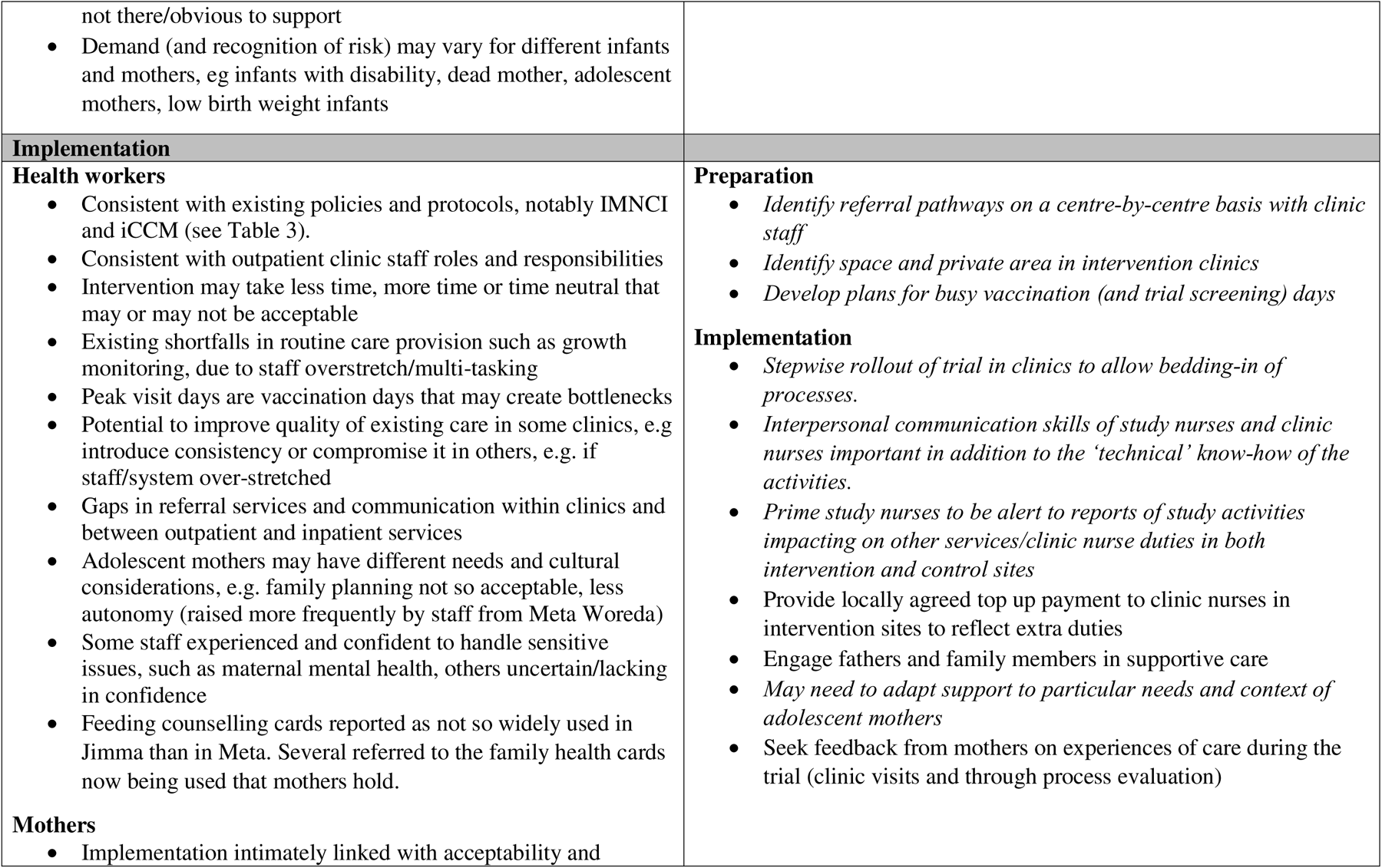
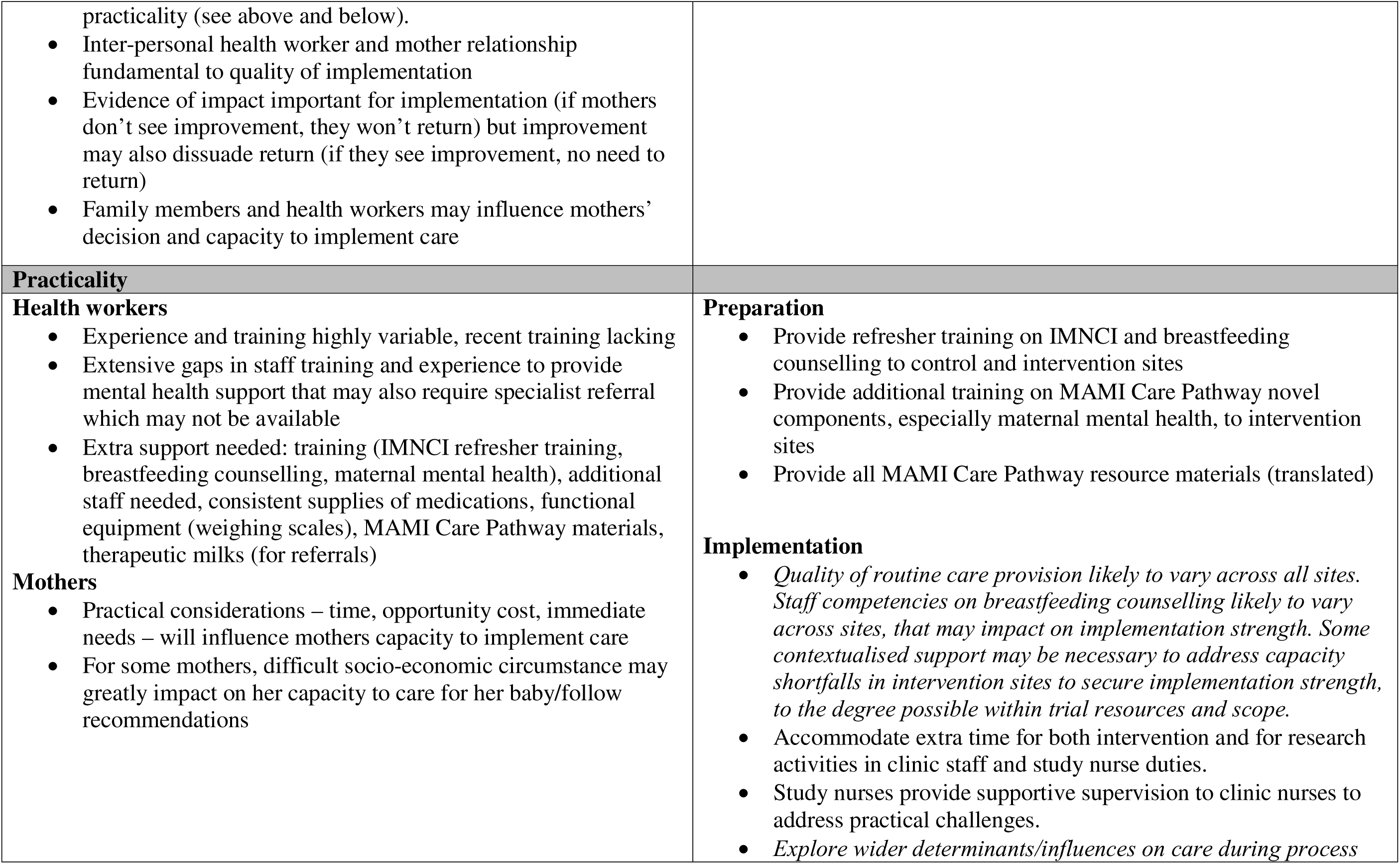
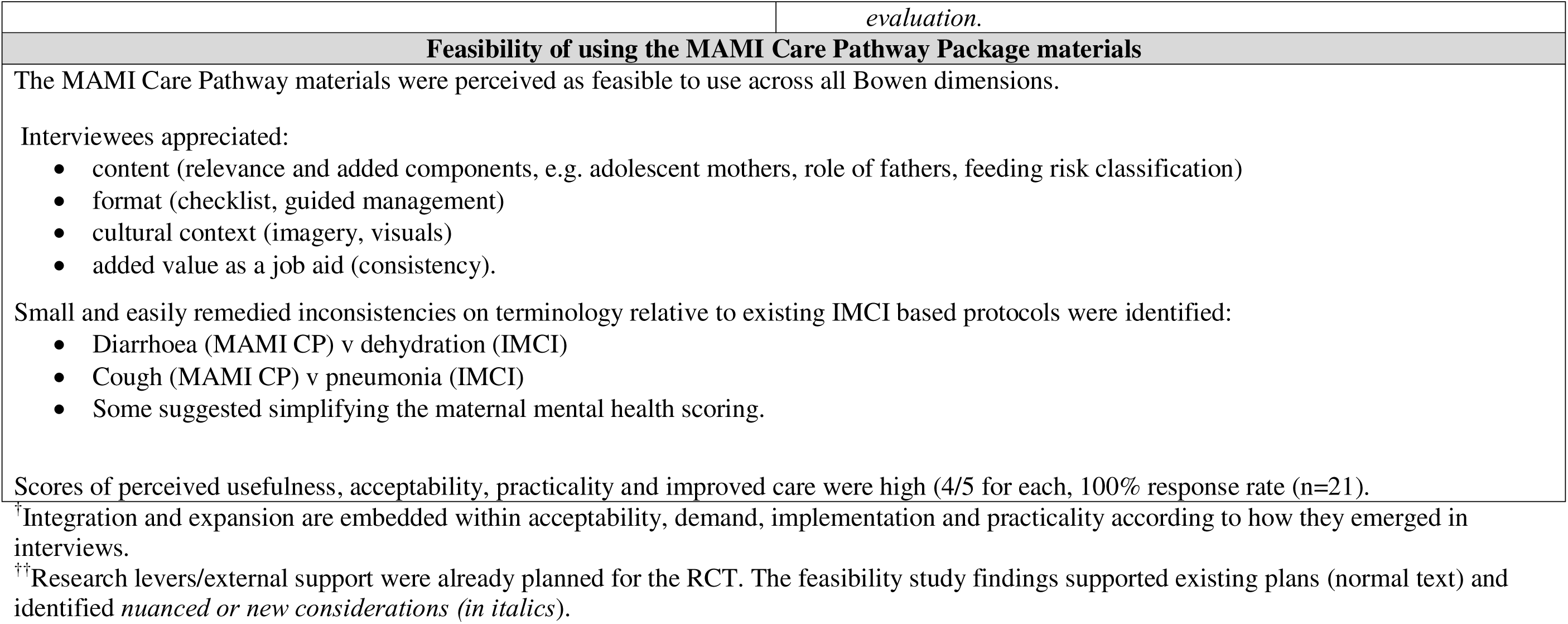
Health workers perceptions of feasibility and implications for RCT preparation and implementation.

#### Acceptability

The MAMI Care Pathway approach was perceived as acceptable to health workers, with some exceptions, for many reasons. Most expressed unacceptability of inpatient severe malnutrition treatment (only current option), evidenced by poor uptake of inpatient referrals by families. One suggested that given consistency of MAMI Care Pathway with government policy, its “*acceptability should not be questioned*”; another felt that policymaker acceptance was a pre-requisite to acceptability by health workers. Suggestions on whom to sensitise on the trial ranged from national policymakers to district managers to community leaders. Several identified key decision-makers who determine service development and change are critical to engage.

Health workers perceived mothers’ acceptability would be influenced by practicalities and previous experiences of care. Evidence of impact of the intervention may improve acceptability for some mothers but may lead to reduced attendance by others once an infant is improving. Many reported that prevalent medication supply stockouts may negatively impact acceptability by fuelling dissatisfaction, undermining the consultation (*“complaining*” mothers) and so reducing attendance.

Health workers expressed that counselling-based interventions may be less acceptable to mothers experiencing food insecurity or economic hardship, especially if commodity-based support is common and expected. This was particularly noted by health workers from Meta, which is a more food insecure area with a history of external (NGO/UN) food assistance. Suggestions of how to improve maternal acceptability (and implementation) of outpatient care included community-tracing for defaulters and provision of social support. Health workers spending time with mothers was also reported (citing experience) to increase their satisfaction and improve acceptability. Health workers noted that interpersonal relationship between themselves and mothers matters, sharing personal examples to illustrate why:

> *If the heath professional approaches a mother in a sociable and compassionate way, the mother may share beyond the health issue. My approach is very friendly and due to this, mothers may tell me something private, like a problem she is having in her relationship with her husband. Sometimes I have called the husband and we have talked together as a family to settle some social issue.*
>
> Clinic nurse, Meta (MHWR001)

Health workers believed that some mothers may accept being asked about their own wellbeing, appreciating its relevance for their baby’s care. For others, this may be unacceptable since too personal; this could cause discomfort and mothers may question this scrutiny. Timing of questions may also influence her acceptance:

> *A mother may be distressed about her child’s health problem and asking about her own condition is not timely. It is better to ask these questions during follow up*.
>
> Health Extension worker, Meta (MHEW001)

Perceived acceptability of sensitive questions on maternal mental health in the context of infant care varied, influenced by acute and chronic mental health status of mothers, cultural context, and health worker relationship. While a health worker may be comfortable to ask the questions, a mother may not be comfortable to answer them. Many expressed that despite the trickiness of asking mental health questions, they were important to investigate as relevant to infant care and “*most of these conditions are not detected unless the mother is asked”.* Some individuals expressed concerns that mental health assessments might lead to over-diagnosing issues, considering that a mother might be acutely stressed when attending with her unwell baby. Several described how they “*put mothers at ease by asking social questions as a gateway*” to more sensitive issues and in *“developing a closer relationship with the mother*” so that she confides in them.

#### Demand

Health workers expressed a strong demand for the MAMI Care Pathway approach to address lack of outpatient care in current acute malnutrition services for infants u6m and to manage less severe cases. They reported experiencing poor job satisfaction and guilt as a result. They wanted help to handle a caseload that they encounter and grapple with *“daily”*. Participants welcomed the proactive combined preventive-treatment approach; waiting until a baby warrants more complicated medical treatment carries greater expense for families and the health system. Some felt the MAMI Care Pathway was needed to help health workers more consistently, comprehensively, and efficiently do their job. Most welcomed embedding maternal care with infant care, especially mental health.

Several health workers expressed the need for outpatient care for at risk infants u6m may not be shared by health service managers, planners and the wider community. Reasons included lack of awareness and misconceptions regarding the problem, prioritisation of services for children over 6 months of age by government, and no routine anthropometric screening. Hence the burden and consequences are “*hidden”* from the formal system that fuels low demand:

> *Whenever there was a very small infant in the family, the family hid the infant at home and do not visit the health facility. Due to this, there is higher chance of the infant dying because of this condition. MAMI Care Pathway implementation safeguards such small infants*.
>
> Health Extension Worker, Meta (MHEW002)

Demand may vary for different infants and mothers, influenced by need, recognition of risk and level of risk, e.g. infants with disability, dead mother, adolescent mothers, and LBW infants. Several health workers expressed a strong need to embed maternal care within infant health services for improved, more efficient, accessible and equitable care:

> *The mother should get services at the same time and contact point as the infant. The MAMI Care Pathway favours integrated care and is appropriate to implement in outpatient care. It is important to help attain equitable health care service delivery.*
>
> District manager, Meta (MMAN001)

#### Implementation

##### Consistency with existing policy and practice

The MAMI Care Pathway was considered highly consistent with existing policy guidance, most citing local IMNCI guidelines that govern outpatient facility care and several referring to iCCM (integrated Community Case Management) that guides community-level care. In practice, they distinguished how IMNCI focuses on “*symptoms and signs of illness*” and *“infant condition and management*”, while the MAMI Care Pathway centres on the mother-infant pair, with “*more detailed assessment and support guidance on feeding, maternal mental health and other MAMI risk factors*” (see further distinctions in Table 3). Most felt the MAMI Care Pathway approach *“fits with staff roles and tasks”* and that “*almost all the activities are what we are performing as routine care”.* The exception was maternal mental health that is not included within IMNCI and ICCM guidance and training packages.

**Table 3:** Distinguishing features of IMNCI compared to MAMI Care Pathway as perceived by health workers.

| **IMNCI** | **MAMI Care Pathway Package** |
| --- | --- |
| Focus on illness/condition | Includes wider risk factors that are preventive/more comprehensive |
| Focus on infant only | Focus on mother-infant pair |
| Distinguishes infants under (<) 2m and 2m-6m | Targets 0 to under 6m as one group |
| Anthropometry: Weight-for-age (applied in growth monitoring) | Anthropometry: weight-for-length (consistent with national malnutrition guidelines), weight for age, MUAC |
| Assesses infant feeding but does not categorise feeding risk | Assesses feeding and categorises feeding risk |
| Includes breastfeeding counselling | Adds additional/targeted content on breastfeeding counselling for risks identified |
| In practice, relies on sick baby presenting to clinic | Uses active screening to identify problems early |
| Does not include maternal mental health | Includes maternal mental health |

Professional satisfaction emerged as an important personal motivator to implement care. Many described how the MAMI Care Pathway could help improve the quality of routine care, which is often done in an *“on-off way”* and “*overlooked”,* by providing practical job aids to guide assessment and management, resulting in more consistent and comprehensive care:

> *In routine care we are working without any protocol which is based on each clinician’s personal experience. This opens inconsistency among the practice of health professionals. The MAMI Care Pathway helps you not to focus on a single isolated health condition and guides you to think in a multi-directional way. Otherwise, you may not think of issues related to feeding and your focus may be on signs and symptoms of illness*.
>
> Clinic nurse, Jimma (JHWR104)

##### Potential for integration and expansion

Integration within IMNCI was proposed as an “*excellent opportunity*” to embed within routine services that could address “*missed opportunities*” to cater for mothers and infants needs in one visit, e.g. screening at vaccination points. Since HEWs are geographically and personally closer to mothers, HEWs were also suggested as having potential to screen for risk, follow up defaulters, and deliver some components of care. “*Awarding educational opportunity, especially for HEWs”* could be a motivator. Such decentralised *“expansion”* would be consistent with government plans and could enable risk-differentiated care:

> *As most of our health centres are going to be scaled-up to second generation level, managing an infant’s condition according to their risk level will be possible in near future*.
>
> District manager, Meta (MMAN001)

However, others cautioned staff and services are already overburdened and underdelivering. Another government initiative, expansion of mandatory health insurance currently being piloted, may increase demand for general health services and have a negative knock-on effect on capacity/supplies for child services.

#### Practicality

##### Extra time

Health worker perspectives of time costs and consequences for health workers and mothers was intimately linked with acceptability and practicality, with much variation and complexity. Some were already informally spending time on these infants, so introduction was considered time neutral:

> *There is no formal guideline. Health workers are working in non-formal ways whenever the client has such problems. I don’t think that implementation will impact the workload of health professionals*.
>
> District manager, Jimma (JMAN002)

Others felt that extra time taken by health workers would be compensated for by better quality of care, greater job satisfaction and decreased burden on inpatient services by preventing malnutrition, saving time (and costs) to the system. Easing health workers in their daily work may further mitigate time issues, making extra time more acceptable, with implementation becoming more efficient over time:

> *When IMNCI began, we thought the package would take longer but it was not the case in practice. When the health workers found the package very helpful, no one was thinking about the time it took. Whenever you start something new, there might be challenges which resolve gradually over time*.
>
> District manager, Jimma (JMAN001)

However, for a few, the time implications meant they perceived the approach was inconsistent with existing roles. Some cautioned that without provision of extra staff time in already stretched services, care may be compromised, especially for more at risk infants:

> *Most infants who visit our health facilities are those who are seriously ill. They need brief assessment and urgent intervention. Taking longer with one infant may affect the care you give to another*.
>
> Clinic nurse, Jimma (JHWR100)

One health worker observed that “*group effort”* and “*commitment”* was needed for implementation. Monitoring and evaluation could help capacitate.

Mothers were perceived to need extra time to attend longer and more frequent appointments, which may or may not be possible. Wider contextual factors beyond the health system were a key influence on practicality. While some mothers may accept that short-term time costs have the potential to avoid future greater time/resource costs, for others this may not be possible (nor acceptable) given family and work commitments. Several referred to *Khat (Catha edulis, a flowering plant native to eastern Africa with psychoactive properties*) trading that is prevalent in Meta and takes up much maternal time (Cochrane & O’Regan, 2016; Habtamu et al., 2023). Others shared examples of mothers working outside the home but still not generating enough income, who would need social assistance and family support to facilitate her care. Several health workers described failed attempts to connect families with the Productive Safety Net Programme (PSNP) (national social protection programme (Abay et al., 2022)) in Meta, with experienced poor child recovery consequences.

##### External support needs

Specific training/supply needs identified by respondents are shown in Table 2. Much of the extra support proposed related to existing shortfalls in routine service implementation, e.g. growth monitoring, referral systems, inter-service information exchange, staffing levels/turnover, and practical constraints, e,g. physical space, shortfalls in supplies, and overcrowded vaccination days. Problems may be potentiated if more components are added without support. Compromised care may reflect pragmatic coping strategies, such as on busy vaccination days:

> *“During vaccination days, health workers just observe the kids physically and if the kid looks emaciated, they link to the health workers working in the under-five clinic for anthropometric measurement and further assessment*.
>
> District manager, Jimma (JMAN002)

Several described insufficient outpatient services for mother’s health, limited to their physical health and at best “*checking for maternal anaemia and screening for malnutrition using Mid Upper Arm Circumference (MUAC)”.* Most identified implementation of the maternal mental health component would be particularly problematic without extra support, given existing policy and service gaps. Some felt the MAMI Care Pathway components would require specialist mental health referrals and was beyond the remit of staff dealing with infants. Others felt that existing clinic staff could be capacitated.

### 3.3 Policymakers’ perspectives

A quantitative summary of the survey responses of senior stakeholders on implementation potential of the MAMI Care Pathway in Ethiopia is included in Supplementary Appendix 2 and qualitative data summarised below.

#### Demand

All participants considered the MAMI Care Pathway approach is needed given the *“infant and child mortality and malnutrition burden in Ethiopia*” that necessitates early identification and intervention, “*shortcomings*” in current guidance and services such as lack of accessible outpatient/home care, variable care quality, disconnected services (e.g. between IMAM and growth monitoring), and gaps in job aids for health workers. At risk infants u6m and mothers were described as “*marginalised*” within guidance and services; “*integrating or mainstreaming*” the approach within existing guidance was needed to address gaps and to avoid other services being *“dropped”* to make way for something new. Inclusion of maternal mental health was described by one MOH respondent as “*very fascinating and promising*”.

#### Consistency

The MAMI Care Pathway aligns with government strategy to treat “*infants in nutrition need*”. It was considered as fully or partially consistent with national policy, respondents referring to IMCNI, iCCM, and guidance on mental health, neonatal care, and IYCF. Identified inconsistencies were not contradictions but reflected additional content covered in the MAMI Care Pathway, such as maternal mental health, embedding maternal and infant care, and child development.

#### Acceptability

Most considered MAMI appropriate and possible to implement *“if adapted to the complex Ethiopian environment”*, in a way that accounts for existing services realities. Some were unsure of its appropriateness for the same reason. Implementation would require increased staff time, dedicated (integrated) service and space, and training with follow up, one noting “*every pilot is possible when capacity is supported”*.

Identified barriers to implementation in outpatient settings related to human resources (staff numbers, competencies), shortfalls in quality of routine care, compromised accessibility for some caregivers (social, geographical) and target-driven health services that may encourage quantity (numbers reached) over quality. Social, service, geographical and cultural contextualisation is needed and expectations of care by users may vary too.

Half felt there were no gaps in the materials, but several cautioned their appraisal was limited by time and expertise. Identified gaps were on performance/evaluation indicators and integration guidance. Several suggested adaptations of existing guidance (IMNCI, iCCM, integrated management of acute malnutrition (IMAM)) to accommodate the MAMI Care Pathway, “*harmonising*” the content that already exists (e.g. feeding assessment) and emphasising the new components, such as mental health (with training). Potential to integrate with iCCM (already at scale through HEWs) would require additional HEW training and/or deployment of extra skilled staff to health posts.

Potential harms (three respondents) were *“at system rather than individual”* level by overstretching staff and service. Identifying workable modifications to address time limitations and first appraising what services (e.g. mental health) are available in different facilities were suggested as mitigating actions.

#### Evidencing policy

Investment in implementation research was suggested as *“critical*” to identify barriers to and cost implications of scale. Most wanted evidence on how to implement the approach within different contexts in Ethiopia, citing affordability, adaptability, feasibility, implementation *“pitfalls”* and how to overcome them, service arrangements, acceptability (by users, by MOH). One suggested to evaluate implementation experiences of existing guidelines.

The planned RCT could help identify what contextual adaptations were needed. Evidence generated through the MAMI RISE Project can influence national policy “*if MOH are involved in every part of the research process”,* whose endorsement is needed to “*activate opportunities for practice”.* Evidence generation could also involve sub-national (regional) and cross-country learning and research.

#### Opportunities

National policy, research or practice opportunities ranged from consistency with government goals (to reduce child mortality, hunger, stunting), government aspirations (to identify cost-effective approaches, comprehensive care provision), and service development plans, such as to upgrade health care provision at selected health posts nationwide. Specific policy opportunities that describe comprehensive care were mentioned (such as the food and nutrition strategy, malnutrition treatment guidelines). Potential to integrate/contribute to updates of guidance on IMNCI, iCCM, maternal and infant and young child feeding, wasting and antenatal care were also identified. Growing attention to early child development and maternal mental health presented an “*opportunity to grab the interest of programme managers and policymakers”*.

## 4.0 Discussion

### Feasibility of MAMI Care Pathway will depend on context

We found the MAMI Care Pathway is likely feasible to implement in outpatient health facilities in Ethiopia, if health workers have additional supports to deliver quality care and informed families have support to access it. Senior stakeholders’ and health workers’ perspectives were consistent in terms of perceived need, policy and service alignment and shortfalls in the current system. Most felt the system could accommodate and benefit from the approach but cautioned limitations and risks to care quality of expanding activities without extra support. Support is needed for existing capacity, upon which the approach depends, and for more ‘innovative’ components, particularly maternal mental health. In some ways, the fact that the MAMI Care Pathway is modelled on what exists is a double-edged sword - it enhances its potential to integrate in existing services, but in practice, is limited by prevailing routine service constraints in Ethiopia (Bayou et al., 2023; Daka, Wordofa, et al., 2023) and elsewhere, with “the policy sacred cow of integrated care repeatedly proving impossible to deliver in practice” (T. Greenhalgh & C. Papoutsi, 2018).

Findings suggest considerable buy-in for the MAMI Care Pathway approach beyond the context of the RCT. Acceptability of health workers appeared driven by a strong demand to address an unmet need for community-based, proactive and preventive care that included mothers’ wellbeing. This was fuelled by dissatisfaction with ‘hidden’ service and personal costs and consequences of shortfalls in accessible care. Some touched on organizational readiness in terms of macro and micro motivation and collective capacity of the system to adopt and sustain the innovation, necessary to facilitate implementation (Domlyn et al., 2021), referring to wider system issues, collective commitment needed and necessary cross-staff, community and government ‘buy-in’. Health workers felt that lack of outpatient services for infants under 6 months reflected poor needs awareness and prioritisation by service developers. However, we found that policymakers were needs aware but service development was hampered by lack of evidence on how to expand care provision and to what effect, within already overburdened services.

Consistent with policymakers, several health workers suggested integration of the approach within IMNCI and iCCM training packages. Similarly, both groups identified potential to expand care provision beyond outpatient clinics, providing some aspects through HEWs that is aligned with government ambitions for more expanded care at selected health post services under the Heath Sector Transformation Plan (HSTP II) (Ministry of Health Ethiopia, 2021). However, given the health extension programme is already “at tipping point”, such expansion might help revitalise but would likely need considerable reform to do so (Zerfu et al., 2023).(Olaghere, 2022)

Many health workers contextualised their perceptions with ‘if’s and but’ qualifiers, reflecting the dynamic and context-specific nature of feasibility within complex systems for health (Moore et al., 2018) (Marten et al., 2022; Skivington et al., 2021). We found the Bowen Framework a helpful tool to unpack the concept of feasibility and guide a deeper exploration. However, strict thematic classification of data was difficult given the fluidity and interdependence we found between themes and potential feedback loops, including between health workers and mothers. For example, the approach was perceived acceptable to some health workers as it helped address an unmet need (*demand*) and was consistent with and supported implementation of routine care *(implementation*) but would require more staff and training to make it workable (*practicality*). Initial ‘open-minded’ deductive analysis helped navigate our Bowen-guided investigation, spotlighting quality of care issues that pervaded Bowen themes; for example, maternal ‘acceptability’ and ‘implementation’ would be affected by mother’s previous experience of services, and whether they are treated sensitively, with respect and dignity; their cultural context and the mother-health worker interpersonal relationship. Using this analytical combination helped to further our understanding on what may shape implementation.

### Research co-creation and partnership strengthens feasibility potential

Involvement of intended users of implementation research should be considered throughout the research process, from identifying priorities, ensuring implementation strategies are relevant, to disseminating and acting on findings (Harvey et al., 2023; Pérez Jolles et al., 2022). Partnerships between researchers and stakeholders are necessary to achieve sound contextual framing of a new intervention, which requires sustained process investment and multi-level brokerage (Partnership Brokers Association, 2019). The strong policy and service alignment we found reflects the nature of the MAMI Care Pathway approach that was collectively developed and modelled on IMCI, and the strong co-creative partnership between the MAMI RISE research partners that regularly consults with intended national and international users. From conceptual stage, planning had future scale in mind (WHO & ExpandNet, 2011), guided by a scaling framework (MSI, 2020) and secured fair allocation of resources across partners (Lavery & IJsselmuiden, 2018). Our findings demonstrate the value of co-creation, partnership and early planning for scale to strengthen contextual fit and future scale potential of complex health interventions (Zamboni et al., 2019) and the potency of combining implicit, contextual and global knowledge to help do so.

### Implications for RCT preparation and implementation

Amongst policymakers and health workers, none questioned whether the proposed approach would work. However, all wanted more evidence (or support) on how to deliver it within existing service realities. These findings resonate with and build on team knowledge and formative work to date (Jibat et al., 2022). They endorse the MAMI RISE approach that combines contextual and global knowledge and experience with contextual implementation research to generate more relevant and transformative implementation knowledge (Michaud-Létourneau et al., 2022), seeking to reduce the evidence-practice/know-do gap that prevails in Ethiopia and globally (Daka, Wordofa, & Woldie, 2023; Harvey et al., 2023).

The value of feasibility testing is recognised (Skivington et al., 2021). However only a quarter of feasibility studies are conducted pre-trial (O’Cathain et al., 2013). Our study evidences the value of investigating implementation context in preparation for an RCT to help connect the science and practice (Harvey et al., 2023) and access critical local knowledge (Abimbola, 2021). Figuring out how to navigate the system during the trial has greatly benefited from a reality-check with those working within it (Reed et al., 2018). Here we share what we learned and how it has informed planning and implementation.

#### Embracing complexity

To generate evidence that improves practice, we need to ‘act scientifically and pragmatically’ whilst ‘embracing the complexity’ of the setting in which change takes place and ‘engaging and empowering’ those responsible for and affected by the change (Reed et al., 2018). In pursuit of this elusive ‘how’, reacting to rather than just reporting on such contextual realities within the trial will be necessary to generate the most practical evidence and insight that policymakers need (Carl R. May et al., 2018). Medical Research Council (MRC) guidance encourages such an approach that varies implementation modality according to context, while maintaining the integrity of the core intervention components (Skivington et al., 2021).

Unpacking the nature and influence of research levers we use, as well as individual and systems reactions and responses during implementation, will be important to capture through RCT evaluation, to identify ‘nugget’ mechanisms of action and any “re-wiring” that happens (Ramani et al., 2022), to contextualise findings, inform transferability and bridge the evidence to practice gap (Harvey et al., 2023).

#### Addressing differing capacity gaps to the degree possible: reality check

The RCT will rely heavily on existing staff competencies. Already planned refresher training was based on assumed competency baseline; our study identified considerable variability and gaps in training and experience of clinic nurses. The degree to which training will address this is likely to vary and will be important to examine during trial implementation. The maternal mental health component was considered both the greatest ‘innovation’ of the approach but the most challenging to implement. Whilst training was already planned, supportive supervision and mentoring during the trial will likely be needed.

We identified factors not modifiable in the context of the RCT that that may impact on implementation fidelity (delivered as intended) (Hasson, 2010) and implementation strength (what it takes to deliver it) (Hargreaves et al., 2016). Embedding mothers’ health as part of infant-focused services is new, acceptable and important but several flagged low prioritisation and limited services to deliver this. RCT consequences will depend on demand, the degree to which they are met and the impact this has on infant outcomes. Mothers in difficult socio-economic circumstance may impede uptake and have implementation consequences, beyond trial influence.

Our findings suggest there will be variability in structural and operational clinic contexts such as space, supplies, referral systems, staff competencies and capacity. Some are more easily modifiable, e.g. allocating space, others less tangible or predictable, e.g. individual staff competencies that will manifest through implementation. Financial/time costs will be compensated to a degree with locally approved ‘top up’ to clinic nurses and provision of transport costs to mothers for trial visits. Study nurses will provide surge support to clinic staff for research activities, and extra staff capacity will be secured where needed.

Our study reinforced that the trial will take time to normalise within research sites. Reflecting this, the RCT developed a three month ‘bedding-in period’ with stepped rollout to clinics, first in Jimma and then in Deder. This enabled more intensive support and ‘learning by doing’ in the initial phase, to inform subsequent clinic rollout.

#### Person-centred care in research in practice

People are at the heart of systems, the “intangible software” that make, shape and break implementation success (Trisha Greenhalgh & Chrysanthi Papoutsi, 2018; Ramani et al., 2022). Our study emphasised that the micro-context of the personal relationship between health workers and mothers may be critical and influential to implementation strength and quality (Carl R. May et al., 2018). Any intervention needs to be sensitive to those human beings at its frontline, to make sure it is a positive experience for them (O’Cathain et al., 2013). This applies to those both receiving care and those delivering it. Our findings emphasise the importance of addressing the inter-personal aspects of care delivery, not just the technical components, as part of training and in on-going support during the trial, especially for more sensitive maternal mental health and for any new staff who will not (yet) have established relationships with mothers. Also, it will be important to watch for and mitigate overstretch of staff that may not only compromise implementation and other services but negatively impact on staff motivation and wellbeing.

#### Capturing the hidden costs of not doing

Health workers touched on the ‘invisible’ costs of not formally dealing with at risk infants and their mothers in services and the ‘savings’ to families and services if they did; these are invisible, hard to measure and so rarely quantified. During the RCT, evaluation of health workers, mothers and fathers’ experiences of care in control sites, will be important to uncover the consequences of ‘not doing’ to help contextualise the more observable ‘price of doing’ in the intervention clinics.

### Making the case for pre-trial feasibility studies

Research levers/external support were already planned for the RCT. The feasibility study findings supported existing plans and identified new or nuanced considerations. Exploring perceived feasibility provided us with depth on areas to strengthen in preparation/implementation and has alerted us to factors within and beyond our control that may impact on implementation fidelity and strength. This has informed initial programme theory development for a realist evaluation to accompany the RCT. Interviewing policymakers engendered interest in the trial (O’Cathain et al., 2013); several policymakers subsequently joining the national RCT Technical Advisory Group. It has sparked us to scope national policy in more detail (McGrath et al., 2023), to most strategically and helpfully contribute to national policy intent and action. We highly recommend feasibility studies precede complex health intervention trials.

### Strengths and limitations

#### Generalisability

As qualitative research, this study is hypothesis generating rather than hypothesis testing; we cannot be sure if our respondents are representative of others, even within Ethiopia. Views are based on preconceptions and past experiences rather than actual experience. Health workers interviewed from Jimma Zone were all currently working in urban health facilities that may have influenced their experiences and perceptions. We did not interview mothers and families directly in our study.

#### Availability

An emergency in Northern Tigray affected availability of some policymakers for survey/interview during data collection (deadlines were extended to help mitigate).

#### Responder bias

Interviewees may have hesitated to challenge the approach that was already planned for trial, and to share their reservations with researchers who were part of the RCT research team. Group orientation pre-interview may have also biased towards a ‘group think’ and positivity. Views were made from a position of safety in their being no personal implications, such as increased workload since those selected would not implement the trial. We did not investigate the perceived feasibility of undertaking the research activities, only the intervention components so perceived time needs are likely underestimates.

### Conclusions

We found that policymakers and health workers perceive the MAMI Care Pathway as feasible to implement during a RCT in outpatient health facilities in Ethiopia, but support will be needed to address prevalent constraints in routine care, capacity gaps for innovative components and to enable sustained access to quality care. Wider contextual influences beyond the influence of the RCT may influence implementation fidelity and strength.

Research co-creation, partnership, early planning for scale and a pre-trial feasibility study has strengthened implementation readiness and scale potential. A responsive RCT that investigates what works and how will generate the most useful evidence for national policymakers.

## Data Availability

All data produced in the present study are available upon reasonable request to the authors.

## Acknowledgements

Thanks to the woreda (district) and clinic staff in Jimma Zone and Meta Woreda and national stakeholders who participated in the interviews. This research was funded by the Eleanor Crook Foundation (MAMI RISE project).

