## Supplementary Appendix 1 for "Strengthening implementation of integrated care for small and nutritionally at-risk infants under six months: pre-trial feasibility study"

### MAMI Care Pathway - who, what, where

Management of small & nutritionally at risk infants under six months & their mothers

#### SCREENING

##### WHAT?

**Screen and refer for assessment using:**

IMCI and MAMI-specific danger signs checks

Infant growth

Infant feeding

Maternal health and wellbeing

##### WHERE?

##### WHO?

##### GUIDES & FORMS

MAMI Rapid Screening Guide

#### ASSESSMENT

##### WHAT?

IMCI and MAMI-specific clinical assessment

Infant growth

Infant feeding

Maternal health and wellbeing

##### WHERE?

##### WHO?

##### GUIDES & FORMS

- MAMI Assessment Guide & Form
- MAMI Feeding Assessment Guide & Form
- MAMI Maternal Mental Health Assessment Guide & Form
- MAMI Enrolment & Follow Up Form

#### SUPPORT AND MANAGEMENT

##### MAMI INPATIENT CARE

###### WHAT?

1. Clinical care to achieve clinical stabilisation.
2. Provide treatment for congenital conditions affecting feeding (e.g. tongue tie), feeding support, and maternal mental health support.
3. If child reaches 6 months of age in inpatient care, conduct 6 month of age outcome review from inpatient facility.

###### WHERE?

###### WHO?

###### GUIDES & FORMS

Refer to existing national guidance on inpatient treatment of infants under six months with wasting and clinical complications

##### MAMI OUTPATIENT CARE

###### WHAT?

1. Counselling on **core topics** for all enrolled pairs.
2. Tailored counselling and actions to address **specific risk factors and problems** as required:
   - Clinical care
   - Feeding counselling & support
   - Mental Health and Psychosocial
   - Support (MHPSS) for mothers
3. Monitoring of progress of mother infant pairs and adjust follow up frequency as appropriate.

###### WHO?

- Clinical care:
- Tailored feeding counselling & support:
- MHPSS for mothers:
- Monitoring of progress:

###### WHERE?

##### REFERRAL TO OTHER AVAILABLE SUPPORT SERVICES AS NEEDED, E.G.

##### GUIDES & FORMS

- MAMI Enrolment & Follow Up Form
- MAMI Counselling Cards & Support Actions Booklet

#### 6-MONTH OF AGE OUTCOME REVIEW

##### WHAT?

Assessment at 6 months of age.

IMCI and MAMI-specific clinical assessment

Infant growth

Infant feeding

Maternal health and wellbeing

Referral to support services for continued care if required.

##### WHERE?

##### WHO?

##### CONTINUUM OF CARE

- Infant malnutrition → wasting treatment
- Feeding problems → IYCF
- Clinical issues → IMCI

##### GUIDES & FORMS

MAMI Enrolment & Follow Up Form

**ACRONYMS** **IYCF –** Infant and Young Child Feeding **MHPSS –** Mental Health and Psychosocial Support **IMCI –** Integrated Management of Childhood Illnesses
