## Supplementary Appendix 2 for "Strengthening implementation of integrated care for small and nutritionally at-risk infants under six months: pre-trial feasibility study"

**Supplementary Appendix 2: Survey responses of senior stakeholders on implementation potential of the MAMI Care Pathway in Ethiopia (n=18)**

| | Yes | | No | | Unsure | | Partially | | With adaption /revision | | Total | |
|---|---|---|---|---|---|---|---|---|---|---|---|---|
| **Survey question** | n | *%* | **n** | *%* | **n** | *%* | **n** | *%* | **n** | *%* | **n** | *%* |
| Is the MAMI Care Pathway Package *consistent* with nutrition and health guidelines in Ethiopia? | 9 | *50%* | 0 | *0%* | 4 | *22%* | 5 | *28%* | *n/a* | *n/a* | 18 | *100%* |
| Is the MAMI Care Pathway *needed* to support care for at-risk infants in outpatient settings? | 18 | *100%* | 0 | *0%* | 0 | *0%* | 0 | *0%* | *n/a* | *n/a* | 18 | *100%* |
| Is the approach *possible* to implement in outpatient setting in Ethiopia? | 3 | *17%* | 0 | *0%* | 1 | *6%* | *n/a* | *n/a* | 14 | *78%* | 18 | *100%* |
| Is the approach *appropriate* to implement in outpatient setting in Ethiopia? | 6 | *33%* | 0 | *0%* | 2 | *11%* | *n/a* | *n/a* | 10 | *56%* | 18 | *100%* |
| Are there *gaps or inconsistencies* in the MAMI Care Pathway Package materials? | 3 | *17%* | 9 | *50%* | 6 | *33%* | *n/a* | *n/a* | *n/a* | *n/a* | 18 | *100%* |
| Do you need *proof or effectiveness evidence* before using the approach in outpatient settings in Ethiopia? | 15 | *83%* | 3 | *17%* | - | - | *n/a* | *n/a* | *n/a* | *n/a* | 18 | *100%* |
| Do you see any *harms* associated with its use in Ethiopia? | 1 | *6%* | 15 | *83%* | 2 | *11%* | *n/a* | *n/a* | *n/a* | *n/a* | 18 | *100%* |
| Do you see any *opportunities* in policy, services or practice to implementation in outpatient settings? | 17 | *94%* | 0 | *0%* | 1 | *6%* | *n/a* | *n/a* | *n/a* | *n/a* | 18 | *100%* |
| Do you see any *barriers* in policy, services or practice to implementation in outpatient settings? | 9 | *50%* | 9 | *50%* | 0 | *0%* | *n/a* | *n/a* | *n/a* | *n/a* | 18 | *100%* |

n/a: not applicable
